# Seroprevalence of anti-SARS-CoV-2 antibodies six months into the vaccination campaign in Geneva, Switzerland

**DOI:** 10.1101/2021.08.12.21261929

**Authors:** Silvia Stringhini, María-Eugenia Zaballa, Nick Pullen, Javier Perez-Saez, Carlos de Mestral, Andrea Loizeau, Julien Lamour, Francesco Pennacchio, Ania Wisniak, Roxane Dumont, Hélène Baysson, Viviane Richard, Elsa Lorthe, Claire Semaani, Jean-François Balavoine, Didier Pittet, Nicolas Vuilleumier, François Chappuis, Omar Kherad, Andrew S. Azman, Klara Posfay-Barbe, Laurent Kaiser, Idris Guessous, the Specchio-COVID19 study group

## Abstract

**Background:** Up-to-date seroprevalence estimates are critical to describe the SARS-CoV-2 immune landscape in the population and guide public health measures. We aimed to estimate the seroprevalence of anti-SARS-CoV-2 antibodies 15 months into the COVID-19 pandemic and six months into the vaccination campaign.

**Methods:** We conducted a population-based cross-sectional serosurvey between June 1 and July 7, 2021, recruiting participants from age- and sex-stratified random samples of the general population. We tested participants for anti-SARS-CoV-2 antibodies targeting the spike (S) or nucleocapsid (N) proteins (Roche Elecsys immunoassays). We estimated the anti-SARS-CoV-2 antibodies seroprevalence following vaccination and/or infection (anti-S antibodies), or infection only (anti-N antibodies).

**Results:** We included 3355 individuals, of which 1814 (54.1%) were women, 697 (20.8%) were aged <18 years and 449 (13.4%) were aged ≥65 years, 2161 (64.4%) tested positive for anti-S antibodies, and 906 (27.0%) tested positive for anti-N antibodies. The total seroprevalence of anti-SARS-CoV-2 antibodies was 66.1% (95% credible interval, 64.1-68.0). We estimated that 29.9% (28.0-31.9) of the population developed antibodies after infection; the rest having developed antibodies only via vaccination. Seroprevalence estimates were similar across sexes, but differed markedly across age groups, being lowest among children aged 0-5 years (20.8% [15.5-26.7]) and highest among older adults aged ≥75 years (93.1% [89.6-96.0]). Seroprevalence of antibodies developed via infection and/or vaccination was higher among participants with a higher educational level.

**Conclusions:** Most adults have developed anti-SARS-CoV-2 antibodies, while most teenagers and children remain vulnerable to infection. As the SARS-CoV-2 Delta variant spreads and vaccination rates stagnate, efforts are needed to address vaccine hesitancy, particularly among younger individuals and socioeconomically disadvantaged groups, and to minimize spread among children.

## Introduction

As the Delta variant (B.1.617.2) of the severe acute respiratory syndrome coronavirus 2 (SARS-CoV-2) drives a surge in new infections worldwide,^1^ vaccination rates stagnate in the USA and much of Europe,^2,3^ undermining public health efforts to achieve herd immunity and curb the pandemic. Up-to-date seroprevalence estimates of anti-SARS-CoV-2 antibodies in the general population remain scarce, yet they are critical in monitoring the progression of infection- and vaccination-induced immune response and informing public health decisions.^4^ The state of Geneva, Switzerland, with a population of about half a million people, has been heavily affected by the pandemic, with 62196 confirmed cases (122 per 1000 inhabitants) and 746 deaths reported by August 11, 2021.^5^ Previous serosurveys of the Geneva population revealed that one in ten individuals had developed anti-SARS-CoV-2 antibodies by April-June 2020 following infection, and only two in ten individuals had done so by November-December 2020,^6^ before mass vaccination began. To date, the only serosurvey conducted after the third wave of the COVID-19 pandemic in the general population reported a total anti-SARS-CoV-2 antibodies seroprevalence of 17.3% in the Portuguese population up to March 2021, including a seroprevalence of 13.1% among self-reported unvaccinated participants.^7^ Using a representative sample of the general population, we aimed to assess the seroprevalence of anti-SARS-CoV-2 antibodies 15 months after the first confirmed case (February 26, 2020) and six months after the vaccination campaign began (December 28, 2020; Pfizer and Moderna vaccines).

## Methods

### Study design and participants

We conducted a cross-sectional serosurvey between June 1 and July 7, 2021, recruiting participants from a random sample of individuals aged 0-64 years provided by the Swiss Federal Office of Statistics, and an age- and sex-stratified random sample of individuals aged 18-24 years and ≥50 years from a previous serosurvey using the same methodology (Addendum S1 in Supplementary Material).^6,8^ The Geneva Cantonal Commission for Research Ethics approved this study (Project N° 2020-00881). Participants gave written consent, provided a venous blood sample, and completed a questionnaire. To detect anti-SARS-CoV-2 antibodies, we used the Roche Elecsys anti-SARS-CoV-2 S and N immunoassays (Roche Diagnostics), both with sensitivity and specificity approaching 100% (Addendum S2-S3).^9^

### Statistical analyses

To estimate seroprevalence (% and 95% credible intervals), we expanded previous Bayesian modelling frameworks that accounted for age, sex, immunoassay performance, and household clustering,^6,8^ to jointly model the antibody response measured by the two immunoassays while additionally accounting for vaccination information. Since the vaccines used to date in Geneva elicit no response to the SARS-CoV-2 N-protein, we used participants’ two-marker antibody profiles to estimate the proportion having any anti-SARS-CoV-2 antibody and the proportion having antibodies due to infection (but could also have been vaccinated). Full details of the statistical model are provided in the supplementary material (Addendum S4). We stratified seroprevalence estimates by sex, age group, and, for ages ≥18 years, education level, and calculated prevalence ratios for sex, age, and education level groups. The model was coded in the probabilistic programming language Stan using the Rstan package^10^ and R version 4.1.^11^

### Role of funding source

The funders had no role in study design, data collection, data analysis, data interpretation, or writing of the report. SS and GP had access to all the data in the study and SS had final responsibility for the decision to submit for publication.

## Results

We included 3355 participants, of whom 1814 (54.1%) were women, 697 (20.8%) were aged <18 years and 449 (13.4%) ≥65 years. Among participants aged ≥18 years, 203 (8.1%) had up to primary education level and 1499 (59.5%) tertiary education level (Table 1).

**Table 1.** Sociodemographic characteristics of sample, serological results, and seroprevalence estimates in Geneva, Switzerland, June 1 to July 7, 2021

|  | Participants<br>N (%) | Vaccinated (self-reported)<sup>c</sup><br>N (%) | Seropositive N (%)<sup>a</sup> |  | Seroprevalence % (95% CrI)<sup>b</sup> |  |
| --- | --- | --- | --- | --- | --- | --- |
|  |  |  | Anti-SARS-CoV-2<br>S protein | Anti-SARS-CoV-2<br>N protein | Antibodies of any<br>origin | Antibodies of infection<br>origin |
| Total | 3355 | 1449 (43.2) | 2161 (64.4) | 906 (27.0) | 66.1 (64.1-68.0) | 29.9 (28.0-31.9) |
| Sex |  |  |  |  |  |  |
| Male | 1541 (45.9) | 669 (43.4) | 995 (64.6) | 416 (27.0) | 65.4 (62.8-68.1) | 30.4 (28.0-33.0) |
| Female | 1814 (54.1) | 780 (43.0) | 1166 (64.3) | 490 (27.0) | 66.7 (64.3-69.1) | 29.5 (27.2-31.9) |
| Age, y |  |  |  |  |  |  |
| 0-5 | 150 (4.5) | 0 (0.0) | 32 (21.3) | 29 (19.3) | 20.8 (15.5-26.7) | 20.8 (15.5-26.7) |
| 6-11 | 281 (8.4) | 0 (0.0) | 100 (35.6) | 95 (33.8) | 31.4 (26.7-36.4) | 31.4 (26.7-36.4) |
| 12-17 | 266 (7.9) | 5 (1.9) | 99 (37.2) | 91 (34.2) | 41.0 (35.6-46.4) | 37.7 (32.5-43.1) |
| 18-24 | 300 (8.9) | 85 (28.3) | 187 (62.3) | 107 (35.7) | 63.6 (57.3-69.8) | 41.8 (36.3-47.5) |
| 25-34 | 372 (11.1) | 121 (32.5) | 212 (57.0) | 97 (26.1) | 61.1 (56.1-65.9) | 31.9 (27.8-36.2) |
| 35-49 | 805 (24.0) | 323 (40.1) | 505 (62.7) | 219 (27.2) | 64.4 (60.2-68.6) | 32.2 (28.7-35.9) |
| 50-64 | 732 (21.8) | 517 (70.6) | 609 (83.2) | 196 (26.8) | 84.7 (81.1-88.1) | 29.8 (26.5-33.4) |
| 65-74 | 207 (6.2) | 174 (84.1) | 187 (90.3) | 41 (19.8) | 89.2 (84.2-93.4) | 22.5 (17.0-28.4) |
| ≥75 | 242 (7.2) | 224 (92.6) | 230 (95.0) | 31 (12.8) | 93.1 (89.6-96.0) | 16.2 (11.8-21.1) |
| Education level<sup>d</sup> |  |  |  |  |  |  |
| Primary | 203 (8.1) | 100 (49.3) | 135 (66.5) | 53 (26.1) | 71.9 (68.4-75.5) | 29.7 (26.6-33.1) |
| Secondary | 818 (32.5) | 393 (48.0) | 560 (68.5) | 219 (26.8) | 70.3 (64.1-76.5) | 27.9 (22.0-33.9) |
| Tertiary | 1499 (59.5) | 878 (58.6) | 1134 (75.7) | 381 (25.4) | 75.2 (72.6-77.9) | 31.7 (28.9-34.4) |

<sup>a</sup> Serology based on Roche Elecsys anti-SARS-CoV-2 S immunoassay and N immunoassay, respectively.
<sup>b</sup> Seroprevalence estimates reported as % and 95% credible interval, adjusted for test performance of both immunoassays and post-stratified to account for age distribution in the Geneva general population and for household clustering of infection and vaccination. N = 3355 for total, and sex- and age-stratified estimates; N = 2520 for education-stratified estimates, excluding participants aged <18 years. Seroprevalence of antibodies of any antibodies is based on proportion of participants with any anti-SARS-CoV-2 antibodies; seroprevalence of infection antibodies is based on proportion of participants who were naturally infected (but could also have been vaccinated).
<sup>c</sup> Self-reported having received at least 1 dose of the COVID-19 vaccine >14 days before blood sample was drawn.
<sup>d</sup> Self-reported education level among participants aged ≥18 years (N = 2520); missing education level values by age groups: n = 15 in 18-24 years group; n = 49 in 25-44 years group; n = 53 in 45-64 years group; n = 2 in 65-74 years group; and n = 19 in ≥75 years group.

Overall, 1449 participants (43.2%) reported receiving at least one COVID-19 vaccine dose ≥14 days before their blood draw, 2161 (64.4%) tested positive for anti-S antibodies, and 906 (27.0%) tested positive for anti-N antibodies (Table 1). The overall seroprevalence estimate was 66.1% (95% CrI 64.1-68.0), including a seroprevalence of 29.9% (28.0-31.9) for infection-derived antibodies (Figure 1). Seroprevalence estimates were similar across sexes, but varied widely across age groups (Table 1 and Figure 1), being lowest among children aged 0-5 years (20.8% [15.5-26.7]) and highest among older adults aged ≥75 years (93.1% [89.6-96.0]). In contrast, the seroprevalence of infection-induced antibodies was lowest among older adults aged ≥75 years (16.2% [11.8-21.1]), and highest among young adults aged 18-24 years (41.8% [36.3-47.5]).

**Figure 1.**
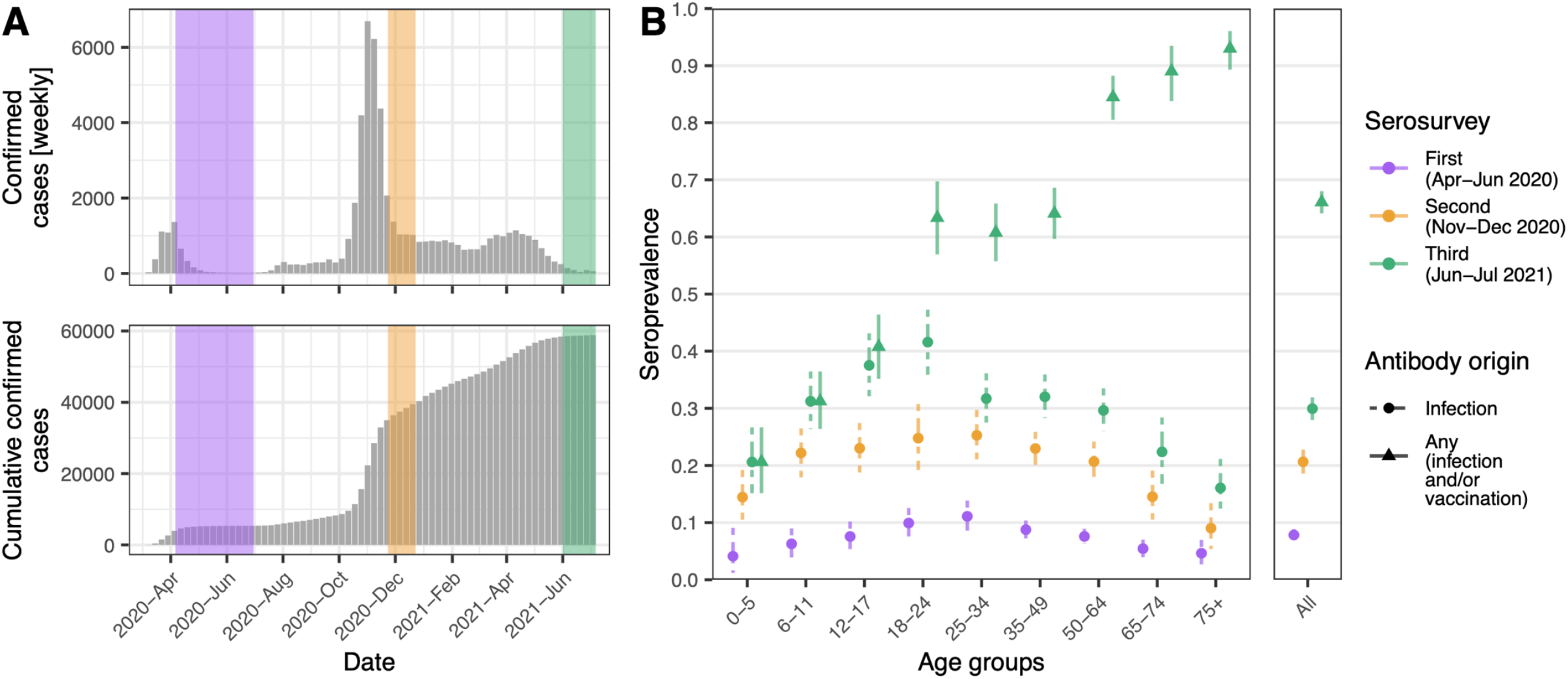
Confirmed SARS-CoV-2 infection cases and estimated seroprevalence of anti-SARS-CoV-2 antibodies in the general population of Geneva, Switzerland, from March 2020 to July 2021 (A) Confirmed SARS-CoV-2 infection cases in the general population of Geneva, Switzerland, from March 2020 to July 2021 (grey; data from https://infocovid.smc.unige.ch/) and serosurvey timings (colored shaded). First serosurvey from April-June 2020 (purple), in which the 0-5 age group only included children aged 5 years ^8^; second serosurvey from November-December 2020 (yellow) ^6^; and third serosurvey from June-July 2021 (green). (B) Seroprevalence estimates (95% credible interval) by age group (in years), serosurvey period, and origin of antibody response.

Among adults with a tertiary education level, 878 (58.6%) reported receiving at least one COVID-19 vaccine dose ≥14 days before their blood draw (Table 1 and Table S1), 1134 (75.7%) tested positive for anti-S antibodies, and 381 (25.4%) tested positive for anti-N antibodies. Among adults with up to a primary education level, 100 (49.3%) reported receiving at least one COVID-19 vaccine dose within the same timeframe, 135 (66.5%) tested positive for anti-S antibodies and 53 (26.1%) tested positive for anti-N antibodies. The overall seroprevalence of antibodies was 75.2 (72.6-77.9) among adult participants with a tertiary education level and 71.9 (68.4-75.5) among those with up to a primary education level (Table 1); the corresponding prevalence ratio for overall antibody seroprevalence was 0.85 (0.77-0.93) among participants with up to primary education relative to those with a tertiary education level (Table S2). The seroprevalence of infection-induced antibodies was similar across education groups.

## Discussion

In this seroprevalence survey, we found that, by July 7, 2021, 66.1% of the population of Geneva, Switzerland, had developed antibodies against SARS-CoV-2 after vaccination and/or infection. We also found that 29.9% of the population had been infected—almost triple the 10.8% seroprevalence reported in April-June, 2020,^8^ and an 8.8 point increase from the 21.1% seroprevalence reported in November-December, 2020,^6^ before the vaccination campaign began (Table S3). This increase in seroprevalence of infection-induced antibodies within a six-month period was largest among young adults aged 18-24 years (16.1 percentage point increase) and teenagers aged 12-17 years (14.1 percentage point increase), suggesting the third COVID-19 wave, comprised primarily of the Alpha (B.1.1.7) variant,^12^ may have particularly affected these age groups (Table S3).

Additionally, we observed marked age differences in seroprevalence, which closely reflect the age-related infection risk observed in Geneva and elsewhere throughout the pandemic^6,8,13^ and, subsequently, the progression of the age-dependent vaccination eligibility in the population since December 2020.^14^ For instance, the 20.8% seroprevalence in children aged 0-5 years—a 5.9 percentage point increase in a six-month period—indicates their consistently reported lower infection risk, and the fact that, to date, vaccination in Switzerland remains approved only for individuals aged ≥12 years. The 16.2% seroprevalence of infection-induced antibodies in adults aged ≥75 years mirrors their previously reported lower infection risk^6,8^ and the fact that they were the first age group targeted by the vaccination campaign; with a reported 92.6% vaccination rate—and an associated estimated total seroprevalence of 93.1%—individuals in this age group were potentially protected from further SARS-CoV-2 infection (Table 1 and Table S3).

We also found that vaccination uptake differed by education level, with a higher proportion of individuals with a tertiary education level reporting being vaccinated than individuals with lower education levels, which reflects socioeconomic inequalities in COVID-19 vaccination reported in the United States and Israel.^15,16^ The proportion having anti-S antibodies was correspondingly higher among individuals with a tertiary education level, though the proportion of anti-N antibodies was similar across education groups, reflecting findings from our previous seroprevalence study,^17^ but differing from patterns of socioeconomic inequalities in infection risk observed in the United Kingdom, Germany, and the United States.^18–20^ While our previous surveys found no socioeconomic inequalities in infection-induced anti-SARS-CoV-2 antibody seroprevalence in the Geneva population,^17^ this study has indicated emerging inequalities in overall seroprevalence, likely driven by the higher vaccination uptake among the more socioeconomically privileged individuals in the population. Indeed, several studies have found a clear pattern of socioeconomic disparities in vaccine hesitancy, whereby a lower proportion of socioeconomically disadvantaged individuals report intention or willingness to get vaccinated against SARS-CoV-2 than more socioeconomically privileged individuals.^21–26^

Strengths of this study include the large sample that is representative of the general population, the measurement of antibodies against both the SARS-CoV-2 S and N proteins, and the robust novel modelling framework accounting for both immunoassays and vaccination status. Limitations include the fact that, as with most seroprevalence surveys,^27^ the sample was generally more socioeconomically advantaged than the general population (Table S4), which may have led to overestimation of vaccine-derived antibody seroprevalence—however, the proportion of vaccinated individuals in our sample was similar to that observed in the general population of Geneva (Table S5); the fact that we only included formal residents and that we only assessed education as a socioeconomic indicator, which may have precluded the identification of inequalities based on other indicators; and, finally, the fact that, since we did not perform neutralization assays, our estimates may not completely reflect SARS-CoV-2 protective immunity.^28^

This study provides the first seroprevalence estimates of SARS-CoV-2 antibodies in a representative sample of the general population after the third pandemic wave and the start of mass vaccination. Our findings highlight how mass vaccination has closed the immunity gap in most of the adult population—particularly among older individuals who are at the greatest risk of severe COVID-19 outcomes.^29^ They attest to the efficacy of free-of-charge vaccination programs in promoting immunization against the virus while highlighting the need to strengthen efforts to address vaccine hesitancy.^21,23^ Importantly, our findings also show that the large majority of children and teenagers, and a large proportion of young and middle-aged adults, lack anti-SARS-CoV-2 antibodies, leaving behind a large reservoir in the population to sustain transmission in the critical months to come.

## Data Availability

Data can be made available upon submission of a data request application to the corresponding author. Virologically-confirmed SARS-CoV-2 infection data are available from the Canton of Geneva: https://infocovid.smc.unige.ch/. Biological material can be reused for further studies upon approval by the cantonal ethics commission of the state of Geneva.

## Contributions

IG, SS, MEZ, NP, JPS, FP, JA, AW, RD, HB, VR, EL, and CS designed study, and acquired data. NP and JPS conducted statistical analyses and created graphics. CdM conducted literature review, wrote the manuscript and created tables. All authors contributed to the interpretation of results and read and approved the final manuscript.

## Declaration of interests

None of the authors have any competing interests to report.

## Data sharing

Data can be made available upon submission of a data request application via the corresponding author. Virologically-confirmed SARS-CoV-2 infection data are available from the Canton of Geneva: https://infocovid.smc.unige.ch/. Biological material can be reused for further studies upon approval by the cantonal ethics commission.

## Acknowledgments

This study would not have been possible without the passionate work of the HEdS students who chose this project for their internship (Marjourie Albarenga, Deborah Amrein, Mayra Battisti, Tatyana Butter, Milica Colovic, Witoria Distinto, Marvorid Jamshedzoda and Nicolas Lechaud), the civil service agents (Vladimir Davidovic and Aniss Moussa), the medical students (Joséphine Duc, Aurélia Hepner, Hugo-Ken Oulevey, Irine Sakvarelidze and Nawel Tounsi), and the caregiver Shamso Hussen. We thank the Hôpital de La Tour and the Clinique de Carouge, as well as the Division of Pediatrics, HUG, for allowing us to use their premises for recruitment. We are deeply grateful to Dr Cyril Sahyoun and his team for our enriching collaboration aimed at easing the experience of children during blood sampling. Finally, we thank all the participants for their interest and invaluable contribution to the study. This study was funded by the Swiss Federal Office of Public Health, the General Directorate of Health of the Department of Safety, Employment and Health of the canton of Geneva, the Private Foundation of the Geneva University Hospitals, the Swiss School of Public Health (Corona Immunitas Research Program) and the Fondation des Grangettes.

## Notes

### Competing Interest Statement

The authors have declared no competing interest.

### Author Declarations

This study was approved by the Geneva Cantonal Commission for Research Ethics (Project Number 2020-00881).

## References

1 Campbell F, Archer B, Laurenson-Schafer H, et al. Increased transmissibility and global spread of SARS-CoV-2 variants of concern as of June 2021. Euro Surveill 2021; 26: 2100509.

2 Diesel J, Sterrett N, Dasgupta S, et al. COVID-19 Vaccination Coverage Among Adults—United States, December 14, 2020–May 22, 2021. Morbidity and Mortality Weekly Report 2021; 70: 922.

3 European Center for Disease Prevention and Control. COVID-19 Vaccine Rollout Report – Week 27. https://covid19-vaccine-report.ecdc.europa.eu/ (accessed July 16, 2021).

4 Murhekar MV, Clapham H. COVID-19 serosurveys for public health decision making. The Lancet Global Health 2021; 9: e559–60.

5 République et Canton de Genève. COVID19 à Genève. Données cantonales. 2021; published online Aug 11. https://infocovid.smc.unige.ch/ (accessed Aug 11, 2021).

6 Stringhini S, Zaballa M-E, Perez-Saez J, et al. Seroprevalence of anti-SARS-CoV-2 antibodies after the second pandemic peak. The Lancet Infectious Diseases 2021; 21: 600–1.

7 Luísa Canto e Castro, Andreia Gomes, Marta Serrano, et al. Longitudinal SARS-CoV-2 seroprevalence in Portugal and antibody maintenance 12 months after the start of the COVID-19 pandemic. Research Square 2021; published online Aug 11. DOI:10.21203/rs.3.rs-603060/v1.

8 Stringhini S, Wisniak A, Piumatti G, et al. Seroprevalence of anti-SARS-CoV-2 IgG antibodies in Geneva, Switzerland (SEROCoV-POP): a population-based study. The Lancet 2020; 396: 313–9.

9 Perez-Saez J, Zaballa M-E, Yerly S, et al. Persistence of anti-sars-cov-2 antibodies: immunoassay heterogeneity and implications for serosurveillance. Clinical Microbiology and Infection DOI:10.1016/j.cmi.2021.06.040.

10 Stan Development Team. Rstan: the R interface to Stan. R package version 2.21.2. 2020. https://mc-stan.org.

11 R Core Team. R: A Language and Environment for Statistical Computing. Vienna, Austria, 2021 https://www.R-project.org/.

12 European Center for Disease Prevention and Control. SARS-CoV-2-increased circulation of variants of concern and vaccine rollout in the EU/EEA, 14th update. 2021 https://www.ecdc.europa.eu/sites/default/files/documents/RRA-15th-update-June=202021.pdf.

13 Bajema KL, Wiegand RE, Cuffe K, et al. Estimated SARS-CoV-2 Seroprevalence in the US as of September 2020. JAMA Internal Medicine 2021; 181: 450–60.

14 Republic and Canton of Geneva. COVID-19 vaccination campaign in Geneva. Republic and Canton of Geneva, Switzerland. https://www.ge.ch/en/node/23804 (accessed July 15, 2021).

15 Barry V, Dasgupta S, Weller DL, et al. Patterns in COVID-19 Vaccination Coverage, by Social Vulnerability and Urbanicity - United States, December 14, 2020-May 1, 2021. MMWR Morb Mortal Wkly Rep 2021; 70: 818–24.

16 Caspi G, Dayan A, Eshal Y, et al. Socioeconomic disparities and COVID-19 vaccination acceptance: a nationwide ecologic study. Clinical Microbiology and Infection 2021; published online June 7. DOI:10.1016/j.cmi.2021.05.030.

17 Richard A, Wisniak A, Perez-Saez J, et al. Seroprevalence of anti-SARS-CoV-2 IgG antibodies, risk factors for infection and associated symptoms in Geneva, Switzerland: a population-based study. medRxiv. 2020; published online Jan 1. http://medrxiv.org/content/early/2020/12/18/2020.12.16.20248180.abstract.

18 Niedzwiedz CL, O’Donnell CA, Jani BD, et al. Ethnic and socioeconomic differences in SARS-CoV-2 infection: prospective cohort study using UK Biobank. BMC medicine 2020; 18: 1–14.

19 Wachtler B, Michalski N, Nowossadeck E, et al. Socioeconomic inequalities in the risk of SARS-CoV-2 infection – First results from an analysis of surveillance data from Germany. Journal of Health Monitoring 2020; : 18–29.

20 Clouston SAP, Natale G, Link BG. Socioeconomic inequalities in the spread of coronavirus-19 in the United States: A examination of the emergence of social inequalities. Social Science & Medicine 2021; 268: 113554.

21 Wisniak A, Baysson H, Pullen N, et al. COVID-19 Vaccination acceptance in the canton of Geneva: A Cross-Sectional Population-Based Study. medRxiv 2021; : 2021.07.05.21260024.

22 Paul E, Steptoe A, Fancourt D. Attitudes towards vaccines and intention to vaccinate against COVID-19: Implications for public health communications. The Lancet Regional Health - Europe 2021; 1: 100012.

23 Sonawane K, Troisi CL, Deshmukh AA. COVID-19 vaccination in the UK: Addressing vaccine hesitancy. The Lancet Regional Health - Europe 2021; 1: 100016.

24 Viswanath K, Bekalu M, Dhawan D, Pinnamaneni R, Lang J, McLoud R. Individual and social determinants of COVID-19 vaccine uptake. BMC Public Health 2021; 21: 818.

25 Rhodes A, Hoq M, Measey M-A, Danchin M. Intention to vaccinate against COVID-19 in Australia. The Lancet Infectious Diseases 2021; 21: e110.

26 Peretti-Watel P, Seror V, Cortaredona S, et al. A future vaccination campaign against COVID-19 at risk of vaccine hesitancy and politicisation. The Lancet Infectious Diseases 2020; 20: 769–70.

27 Accorsi EK, Qiu X, Rumpler E, et al. How to detect and reduce potential sources of biases in studies of SARS-CoV-2 and COVID-19. European Journal of Epidemiology 2021; 36: 179–96.

28 Chia WN, Zhu F, Ong SWX, et al. Dynamics of SARS-CoV-2 neutralising antibody responses and duration of immunity: a longitudinal study. The Lancet Microbe 2021; 2: e240–9.

29 Bonanad C, García-Blas S, Tarazona-Santabalbina F, et al. The Effect of Age on Mortality in Patients With COVID-19: A Meta-Analysis With 611,583 Subjects. Journal of the American Medical Directors Association 2020; 21: 915–8.

