## Supplemental material for "Seroprevalence of anti-SARS-CoV-2 antibodies six months into the vaccination campaign in Geneva, Switzerland"

**Addendum S1. Specchio-COVID-19 study group**

Isabelle Arm-Vernez, Andrew S Azman, Fatim Ba, Oumar Ba, Delphine Bachmann, Jean-François Balavoine, Michael Balavoine, Hélène Baysson, Lison Beigbeder, Julie Berthelot, Patrick Bleich, Gaëlle Bryand, François Chappuis, Prune Collombet, Delphine Courvoisier, Alain Cudet, Carlos de Mestral, Paola D'ippolito, Richard Dubos, Roxane Dumont, Isabella Eckerle, Nacira El Merjani, Antoine Flahault, Natalie Francioli, Marion Frangville, Idris Guessous, Séverine Harnal, Samia Hurst, Laurent Kaiser, Omar Kherad, Julien Lamour, Pierre Lescuyer, François L'Huissier, Fanny-Blanche Lombard, Andrea Jutta Loizeau, Elsa Lorthe, Chantal Martinez, Lucie Ménard, Lakshmi Menon, Ludovic Metral-Boffod, Benjamin Meyer, Alexandre Moulin, Mayssam Nehme, Natacha Noël, Francesco Pennacchio, Javier Perez-Saez, Giovanni Piumatti, Didier Pittet, Jane Portier, Klara M Posfay-Barbe, Géraldine Poulain, Caroline Pugin, Nick Pullen, Zo Francia Randrianandrasana, Aude Richard, Viviane Richard, Frederic Rinaldi, Jessica Rizzo, Khadija Samir, Claire Semaani, Silvia Stringhini, Stéphanie Testini, Didier Trono, Guillemette Violot, Nicolas Vuilleumier, Ania Wisniak, Sabine Yerly, María-Eugenia Zaballa

**Addendum S2. Recruitment of participants**

The study sample was built from a combination of newly recruited individuals, randomly selected from official population registers provided by the Swiss Federal Statistics Office, and participants from previous population-representative seroprevalence surveys [1,2] (eFigure 1). Newly selected individuals (aged 0 to 64 years) were invited by letter, while returning participants (≥18 years) were invited by letter or email when available. Individuals not having responded to the first invitation received up to two written reminders and were contacted by phone when contact numbers were available. Children and teenagers (<18 years) were invited to come with the members of their household, adult participants received individual invitation. Participation rates differed between age groups and depending on previous participation, ranging from 18.9% for newly-recruited children aged <6 years to 80.0% for returning participants aged ≥65 years. A comparison of age and sex distribution of study sample and the Geneva population is provided in eFigure 2.

**Addendum S3. Immunoassays**

We used two commercially-available immunoassays to assess anti-SARS-CoV-2 antibodies; the Roche Elecsys anti-SARS-CoV-2 S immunoassay, which detects immunoglobulins (IgG/A/M) against the receptor binding domain of the virus spike (S) protein (#09 289 275 190, Roche-S), and has an in-house sensitivity of 99.6% (95% CrI, 98.3%-100%) and specificity of 99.8% (95% CI, 99.3%-100%)[3,4]; and the Roche Elecsys anti-SARS-CoV-2 N immunoassay, which detects immunoglobulins (IgG/A/M) targeting the virus nucleocapsid (N) protein (#09 203 079 190, Roche-N), and has an in-house sensitivity of 99.8% (95% CrI, 99.4%-100%) and specificity of 99.1% (95% CI, 98.3%-99.7%)[3] (Roche Diagnostics, Rotkreuz, Switzerland). We defined seropositivity using the manufacturer's provided cut-off value of titer ≥0.8 U/mL for the Roche–S, and cut-off index ≥1.0 for the Roche-N immunoassays. Anti-SARS-CoV-2 antibodies were detected in all but three participants reporting having received at least one dose of the COVID-19 vaccine more than 14 days before serological assessment.

### Addendum S4. Overview of Statistical Framework

Our aim was to infer the proportion of the population having any antibody against SARS-CoV-2, as well as the proportion of those who acquired antibodies through natural infection as opposed to vaccination. We do so by modelling jointly the antibody response measured by the Roche-N and Roche-S immunoassays together with participants' responses to a vaccination questionnaire. We disentangle natural infection from vaccination antibody responses using the fact that the only available vaccines in Switzerland to date—the mRNA-1273 from Moderna/US NIAID,[5] and the mRNA-BNT162b2/Comirnaty from Pfizer/BioNTech[6]—both elicit a response exclusively to the S protein of SARS-CoV-2, as opposed to natural infections which typically elicit a response to both the N and S virus proteins. We expand previous Bayesian modelling frameworks used for seroprevalence estimates that account for demographic parameters (sex and age), test performance and household infection clustering.[1,2] The main additions to the previous models are that we now model jointly the response to both tests, and that we account for vaccination-induced antibody response.

#### *S4.1 Multinomial response model*

We model the Roche-S and Roche-N tests results for participant $i$, $x_i$, consisting of one of four possible outcome combinations $[n_{S^+N^+}, n_{S^-N^+}, n_{S^+N^-}, n_{S^-N^-}]$ (+ indicates antibody presence; $-$ indicates absence) with $x_i \in \{[1,0,0,0],[0,1,0,0],[0,0,1,0],[0,0,0,1]\}$ using a multinomial distribution parametized by parameter vector $\pi_i = [\pi_i^{++}, \pi_i^{-+}, \pi_i^{+-}, \pi_i^{--}]$, where $\pi^{lm}$ is the probability of having Roche-S test result $l$ and Roche-N result $m$, accounting both for the underlying probability of each antibody status $p^{\pm/\pm}$, and test sensitivity, $\theta^+$and specificity, $\theta^-$:

$$x_i \sim \text{Multinomial}(\pi_i)$$

$$\pi_i^{++} = \theta_S^+\theta_N^+p_i^{++} + (1-\theta_S^-)\theta_N^+p_i^{-+} + \theta_S^+(1-\theta_N^-)p_i^{+-} + (1-\theta_S^-)(1-\theta_N^-)p_i^{--},$$

$$\pi_i^{-+} = (1-\theta_S^+)\theta_N^+p_i^{++} + \theta_S^-\theta_N^+p_i^{-+} + (1-\theta_S^+)(1-\theta_N^-)p_i^{+-} + \theta_S^-(1-\theta_N^-)p_i^{--},$$

$$\pi_i^{+-} = \theta_S^+(1-\theta_N^+)p_i^{++} + (1-\theta_S^-)(1-\theta_N^+)p_i^{-+} + \theta_S^+\theta_N^-p_i^{+-} + \theta_S^-(1-\theta_N^-)p_i^{--},$$

$$\pi_i^{--} = (1-\theta_S^+)(1-\theta_N^+)p_i^{++} + \theta_S^-(1-\theta_N^+)p_i^{-+} + (1-\theta_S^+)\theta_N^-p_i^{+-} + \theta_S^-\theta_N^-p_i^{--}.$$

The underlying probability of antibody status accounts both for the probability of natural infection $\leftthreetimes_i$ and vaccination status, $v_i \in \{0,1\}$. Following our previous modeling frameworks,[1,2] we model the probability of natural infection as a function of sex and age category, accounting for household infection clustering through a random effect, $\alpha_h$:

$$logit(\leftthreetimes_i) = \alpha_h + \mathbf{X}_{\boldsymbol{i}}\boldsymbol{\beta}$$

$$\alpha_h \sim Normal(0, \sigma_h^2)\ ,$$

where $\mathbf{X}_i$ is the matrix of covariates, and $\boldsymbol{\beta}$ the vector of regression coefficients. The probabilities of antibody status are then given by:

$$p_i^{++} = \gamma^{++}\leftthreetimes_i + \gamma^{-+}v_i$$

$$p_i^{-+} = \gamma^{-+}\leftthreetimes_i\ (1-v_i)$$

$$p_i^{+-} = (1-\leftthreetimes_i)v_i + \gamma^{+-}\leftthreetimes_i$$

$$p_i^{--} = 1 - v_i(1-\leftthreetimes_i) - \leftthreetimes_i,$$

where $\gamma^{++}, \gamma^{-+}, \gamma^{+-}$ are the conditional probability of having $S^+N^+$, $S^-N^+$, $S^+N^-$ responses, respectively, upon natural infection, $v_i$ is the probability of having a vaccine-induced $S^+$ response as a function of the conditional probability of antibody response upon infection $\eta_i$, $v_i = \eta_i \times v_i$.

#### *S4.2 Vaccination*

To obtain population-level seroprevalence estimates, we also model the proportion of vaccinated individuals in each sex/age class following the approach used for natural infection:

$$v_i \sim \text{Bernoulli}(\phi_i)$$

$$logit(\phi_i) = \alpha_{v,h} + \mathbf{X}_i\boldsymbol{\beta}_{\boldsymbol{v}}$$

$\alpha_{v,h} \sim \text{Normal}(0, \sigma_v^2)$

Given vaccination policy recommendations in the state of Geneva, previously infected individuals were discouraged from being vaccinated in the early phase of the vaccination campaign, thus making the probability of vaccination dependent on the infection status of the individual. We account for this dependence by modelling separately the probability of vaccination given the infection status and marginalizing out the infection status:

$$P(v_i|\mathbf{\Theta}) = \text{Bernoulli}(v_i|\phi_i^I) \leftthreetimes_i + \text{Bernoulli}(v_i|\phi_i^{\sim I})(1 - \leftthreetimes_i),$$

$$logit(\phi_i^{\sim I}) = \alpha_{v,h} + \mathbf{X}_i \boldsymbol{\beta}_{\boldsymbol{v}},$$

$$logit(\phi_i^I) = \alpha_{v,h} + \mathbf{X}_i \boldsymbol{\beta}_{\boldsymbol{v}} + \mathbf{X}_i \boldsymbol{\beta}_{\boldsymbol{v}}^{\boldsymbol{I}}$$

$$\alpha_{v,h} \sim \text{Normal}(0, \sigma_v^2),$$

where $\mathbf{\Theta}$ is the vector of all model parameters, $I, \sim I$ indicates infection and non-infection, respectively, and $\boldsymbol{\beta}_{\boldsymbol{v}}^{\boldsymbol{I}}$ is the vector of regression coefficients giving the difference in probability of vaccination between infected and non-infected individuals.

When estimating the population-level seroprevalence, we account for the conditional probability of vaccination given infection, $p_{v|\sim}$, in the probability of a negative S and N response accounting for household vaccination clustering, $p^{--}$, as:

$$p_{s,k}^{--} = 1 - p_{v|\sim I,s,k} \times \left(1 - p_{I,s,k}\right) - p_{I,s,k},$$

where $s, k$ denote the sex and age categories, $p_{v|\sim I,s,k} = \int_0^1 \emptyset_{s,k}^{\sim I}(t)dt = \int_0^1 \beta_{v,s,k} X_{s,k} + \sigma_v \Phi^{-1}(t)dt$, with $\Phi^{-1}(t)$ being the normal quantile function, and similarly, $p_{I,s,k}$ is the probability of infection with $p_{I,s,k} = \int_0^1 \leftthreetimes_{s,k}(t)dt = \int_0^1 \beta_{s,k} X_{s,k} + \sigma \Phi^{-1}(t)dt$.

#### *S4.3 Diagnostic test performance*

The individual performance of both N and S tests is incorporated hierarchically following Gelan & Carpenter.[7] The sensitivity, $\theta^+$, is determined using $n^+$ RT-PCR positive controls from a laboratory validation study,[8] of which $x^+$ tested positive. The specificity, $\theta^-$, is determined using $n^-$ pre-pandemic negative controls, of which $x^-$ tested positive. For the Roche N test, these values are modulated by data in Ainsworth et al.[9] For the Roche S test, the laboratory study data are modulated by those available on the Roche website (last accessed July 19, 2021).

#### *S4.4 Priors*

We follow a similar setting of the priors on the tests' sensitivity and specificity as Gelman & Carpenter.[7] For the study , the specificity $\theta_j^-$ and sensitivity $\theta_j^+$ are drawn from normal distributions on the log odds scale:

$$logit\left(\theta_j^-\right) \sim \text{Normal}(\mu_{\theta-}, \sigma_{\theta-})$$

$$logit\left(\theta_j^+\right) \sim \text{Normal}(\mu_{\theta+}, \sigma_{\theta+}).$$

Hyperparameters $\mu_z$ and $\sigma_z$ for $z \in (\theta^-, \theta^+)$ follow, on the logit scale, normal distributions $\mu_z \sim \mathcal{N}(4,2)$ and positive half-normals $\sigma_z \sim \mathcal{N}^+(0,1)$, respectively. These priors on test performance were identical for both the Roche S and Roche N tests.

We used standard normal $\mathcal{N}(0,1)$ priors for the logistic regression coefficients for infection $\boldsymbol{\beta}$. For coefficients of vaccination $\boldsymbol{\beta}_{\boldsymbol{v}}$ and for coefficients of the difference in probability of vaccination between infected and non-infected individuals $\boldsymbol{\beta}_{\boldsymbol{v}}^{\boldsymbol{I}}$, we also used standard normal except for the youngest age group (ages 0-5 years and 6-11 years). For these two age groups, $\beta_v \sim \mathcal{N}(-10, 0.01)$ to reflect the fact that there was almost no vaccination in these youngest age groups in Geneva at the time of the study (NB vaccination registration for those aged 12-15 years opened on June 16, 2021: https://www.ge.ch/en/getting-vaccinated-against-covid-19/covid-19-vaccination-campaign-geneva, last accessed July 20, 2021). Correspondingly, $\beta_v^I \sim \mathcal{N}(0, 0.01)$ for these two age groups.

The priors for the means of the household random effects $\alpha_h$ and $\alpha_{h,v}$, followed standard normal, and for standard deviations of the household random effects were positive half-normals, $\sigma_h \sim \mathcal{N}^+(0, 2)$ and $\sigma_v \sim \mathcal{N}^+(0, 2)$. We use a Dirichlet prior on the conditional probability of having $S^+N^+$, $S^-N^+$, $S^+N^-$ responses upon natural infection, $\gamma^{++}, \gamma^{-+}, \gamma^{+-}, \gamma \sim \text{Dir}(10, 1, 1)$, to highly favour production of both anti-S and anti-N antibodies upon infection. Finally, we put a strong prior on the conditional probability of antibody response after vaccination $\eta_i \sim \text{Beta}(10, 0.1)$.

***S4.5 Implementation***

The model was coded in the probabilistic programming language Stan[10] using the Rstan package.[11] R[12] version 4.1 was used for data analysis. Four chains were run with 1500 iterations each, 250 of which were warmup, to give a total of 5000 posterior samples. Convergence was assessed by checking that $\widehat{R} \approx 1$, that the effective sample size was reasonable for all parameters, and visually using shinystan[13] diagnostics checks.

**Figure S1. Participants recruitment and inclusion into analytical sample**

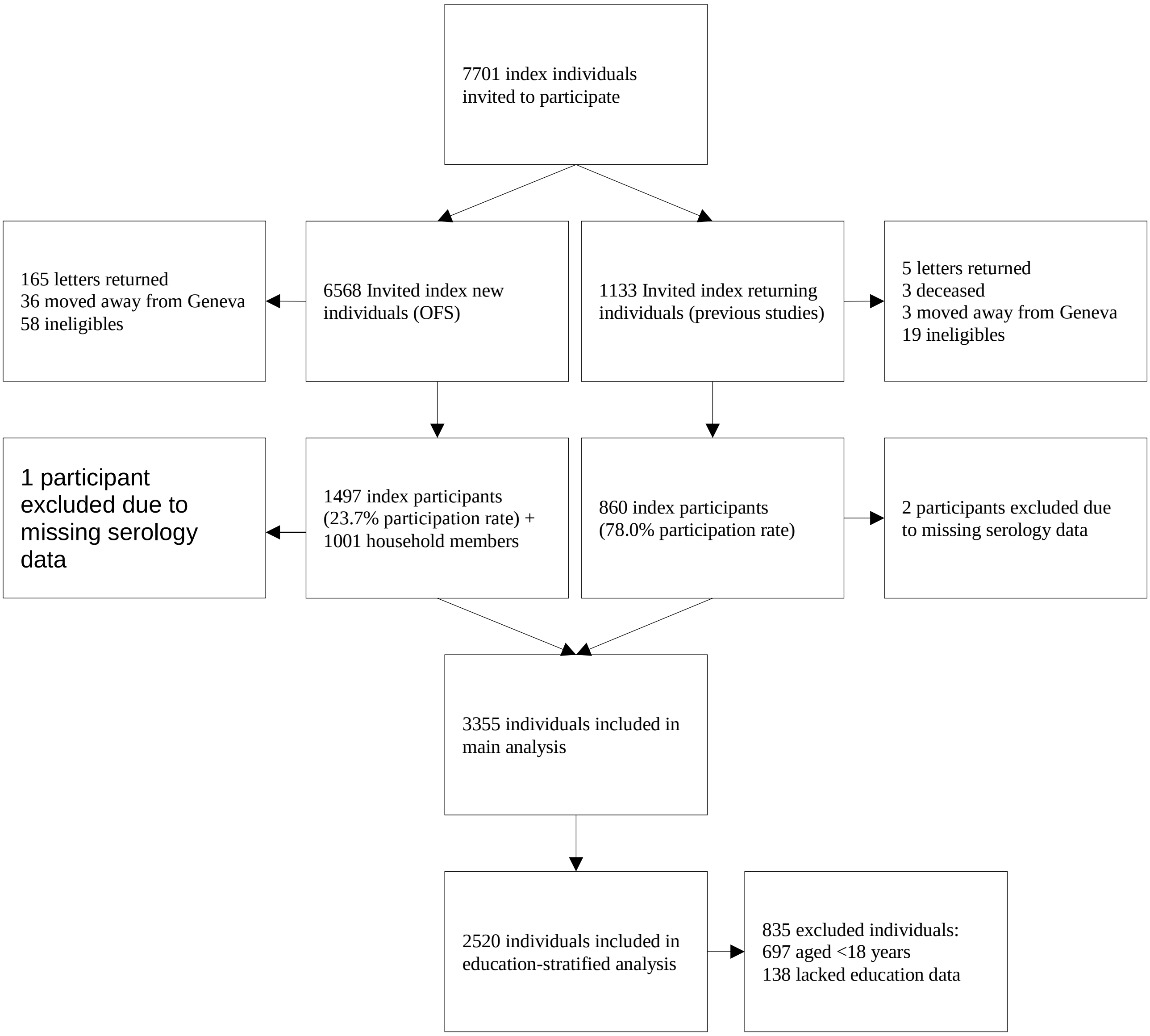

**Figure S2. Comparison of age and sex composition of study sample (bars) and the Geneva population (dots)**

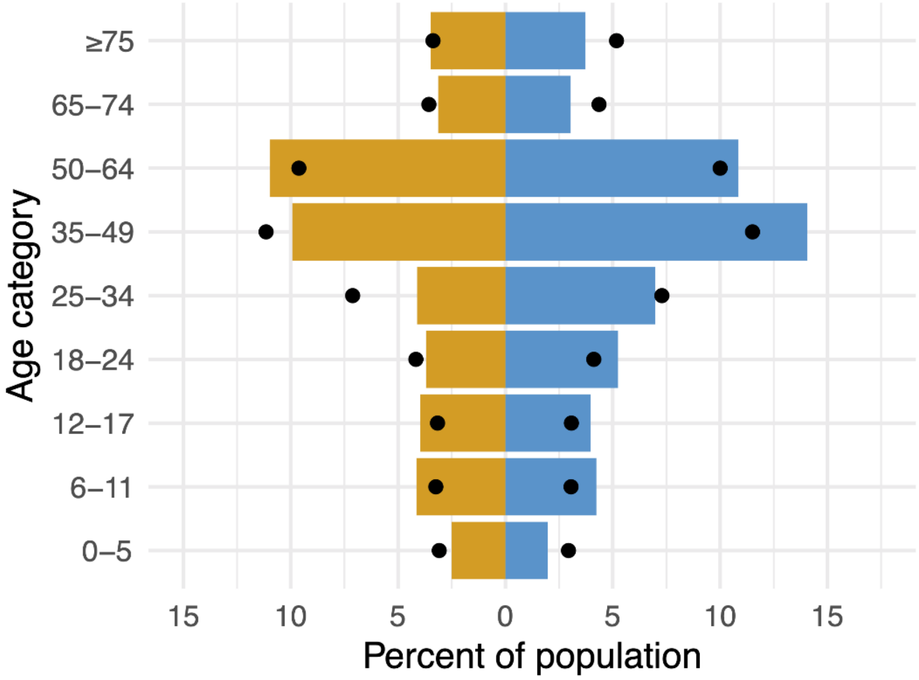


Dark yellow represents males; blue represents females

**Figure S3. Quantitative values of Roche-S and Roche-N immunoassays results**

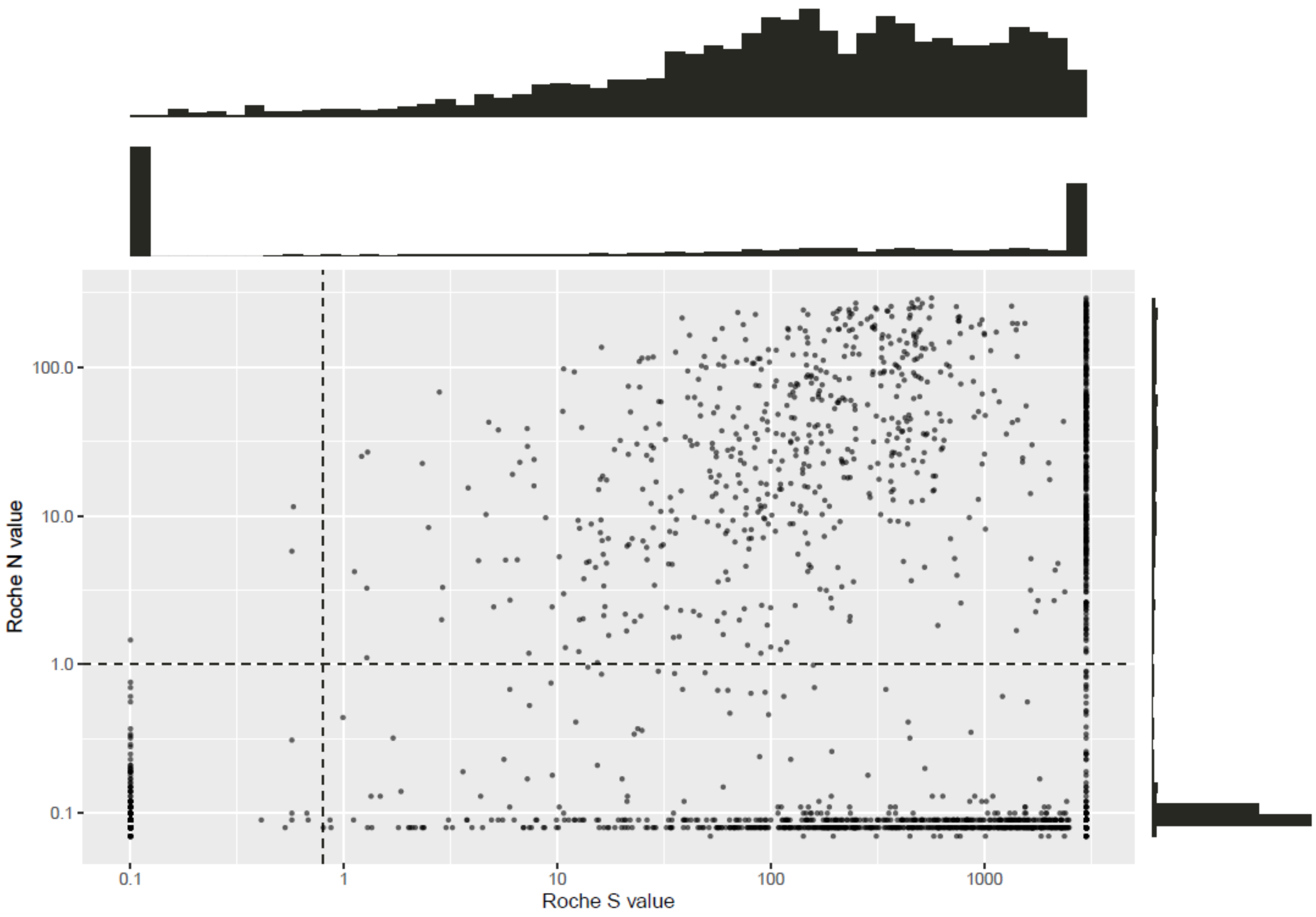


Each dot represents one participant. N and S value units are U/mL. For Roche-S, any values <0.4 were coded as 0.1, and any >2500 were coded as 3000 for ease of viewing, as our lab results do not provide more detailed data. The upper histogram for Roche S is thus a histogram without these 2 extremes i.e. binned from 0.4 to 2500 U/mL.

**Figure S4. Antibodies response category and vaccination status**

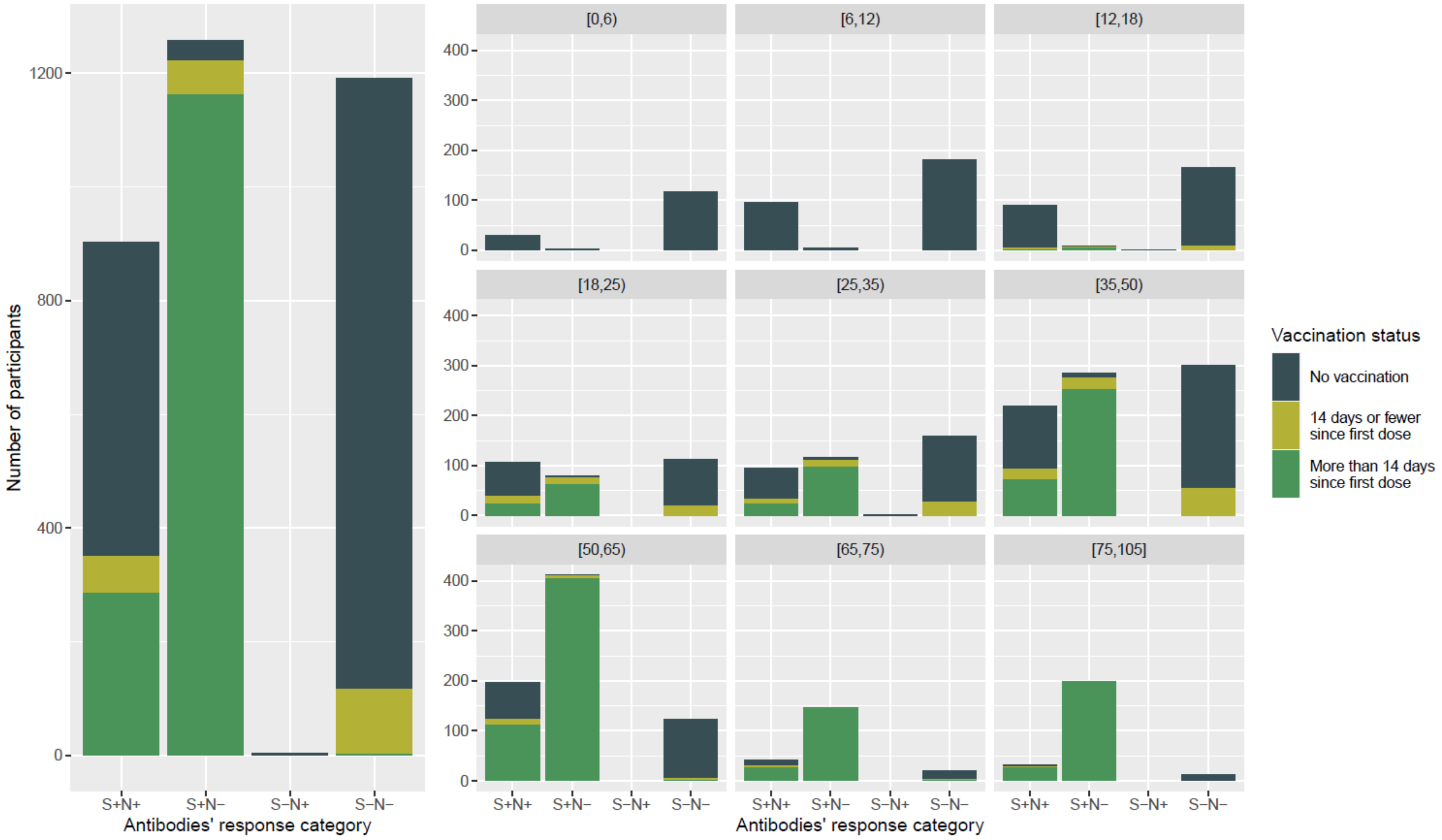


Number of participants in the four possible categories of the S and N tests. + indicates antibodies detected; - indicates antibodies not detected.

**Table S1. Proportion of participants having received at least 1 dose of the COVID-19 vaccine more than 14 days before serological assessment.**

| | Participants | Vaccinated, at least one dose | |
|---|---|---|---|
| | N (%) | Reported[a] N (%) | Estimated[b] % (95% CrI) |
| Total | 3355 | 1449 (43.2) | 44.9 (43.4-46.4) |
| Sex | | | |
| Male | 1541 | 669 (43.4) | 43.4 (41.3-45.5) |
| Female | 1812 | 780 (43.0) | 46.3 (44.5-48.1) |
| Age, y | | | |
| 0-5 | 150 | 0 (0.0) | 0.0 (0.0-0.0) |
| 6-11 | 281 | 0 (0.0) | 0.0 (0.0-0.0) |
| 12-17 | 266 | 5 (1.9) | 3.7 (1.9-6.2) |
| 18-24 | 300 | 85 (28.3) | 28.6 (23.8-33.6) |
| 25-34 | 372 | 121 (32.5) | 35.6 (31.1-40.2) |
| 35-49 | 805 | 323 (40.1) | 40.9 (37.3-44.5) |
| 50-64 | 732 | 517 (70.6) | 70.1 (66.7-73.3) |
| 65-74 | 207 | 174 (84.1) | 81.5 (75.8-86.5) |
| ≥75 | 242 | 224 (92.6) | 90.0 (86.0-93.5) |
| Education level[c] | | | |
| Primary | 203 | 100 (49.3) | 51.5 (48.5-54.4) |
| Secondary | 818 | 393 (48.0) | 52.9 (46.8-58.8) |
| Tertiary | 1499 | 878 (58.6) | 56.0 (53.6-58.4) |

[a] Self-reported having received at least one dose of any COVID-19 vaccine, more than 14 days before blood drawing.

[b] Estimated vaccinated proportion in population, reported as % and 95% credible interval, adjusted for test performance of both immunoassays and post-stratified to account for age distribution in the Geneva general population and for household clustering of infection and vaccination.

[c] Self-reported education level among participants aged ≥18 years (N = 2520).

**Table S2. Prevalence ratio for seroprevalence of anti-SARS-CoV-2 antibodies in Geneva, Switzerland, from June 1 to July 7, 2021**

| | | Prevalence ratio (95% CrI)[a] | | | |
|---|---|---|---|---|---|
| | **Participants N** | **Antibodies of any origin** | ***P value*** | **Antibodies of infection origin** | ***P value*** |
| Sex | | | | | |
| Male | 1541 | 1.01 (0.94-1.08) | 0.75 | 1.01 (0.92-1.11) | 0.81 |
| Female | 1814 | 1.00 (ref.) | | 1.00 (ref.) | |
| Age, y | | | | | |
| 0-5 | 150 | 0.34 (0.25-0.44) | <.0001 | 0.65 (0.48-0.85) | 0.002 |
| 6-11 | 281 | 0.52 (0.43-0.61) | <.0001 | 0.99 (0.81-1.20) | 0.86 |
| 12-17 | 266 | 0.67 (0.57-0.78) | <.0001 | 1.19 (0.98-1.42) | 0.08 |
| 18-24 | 300 | 1.04 (0.92-1.17) | 0.49 | 1.32 (1.10-1.57) | 0.003 |
| 25-34 | 372 | 1.00 (ref.) | | 1.00 (ref.) | |
| 35-49 | 805 | 1.06 (0.96-1.16) | 0.27 | 1.01 (0.86-1.19) | 0.89 |
| 50-64 | 732 | 1.39 (1.28-1.52) | <0.0001 | 0.94 (0.79-1.10) | 0.43 |
| 65-74 | 207 | 1.47 (1.34-1.60) | <0.0001 | 0.71 (0.52-0.92) | 0.01 |
| ≥75 | 242 | 1.53 (1.41-1.67) | <0.0001 | 0.51 (0.36-0.68) | <0.0001 |
| Education level[b] | | | | | |
| Primary | 203 | 0.85 (0.77-0.93) | <0.0001 | 0.93 (0.81-1.07) | 0.28 |
| Secondary | 818 | 0.84 (0.72-0.96) | 0.01 | 0.88 (0.69-1.08) | 0.22 |
| Tertiary | 1499 | 1.00 (ref.) | | 1.00 (ref.) | |

[a] Prevalence ratio (95% credible interval) from Bayesian multinomial regression models accounting for sex, age, test performance and household clustering. Reference group for age and sex estimates (N = 3355) is female, ages 24-35 years. For education level estimates (N =2520), reference group is female, ages 25-44 years with tertiary education level.

[b] Self-reported education level among participants aged ≥18 years (N = 2520).

**Table S3. Comparison of seroprevalence of antibodies[a] naturally developed through infection by November-December 2020 and June-July 2021, Geneva, Switzerland**

| | Geneva population N | Seroprevalence of antibodies after infection % | | Percent increase | Absolute increase % points | Absolute increase N |
|---|---|---|---|---|---|---|
| | | Nov-Dec 2020 | Jun-Jul 2021 | | | |
| Total | 508774 | 21.1 | 29.9 | 42% | 8.8 | 44772 |
| Sex | | | | | | |
| Male | 246655 | 21.9 | 30.4 | 39% | 8.5 | 20966 |
| Female | 262119 | 20.4 | 29.5 | 45% | 9.1 | 23853 |
| Age, y | | | | | | |
| 0-5 | 30633 | 14.9 | 20.8 | 40% | 5.9 | 1807 |
| 6-11 | 32041 | 22.8 | 31.4 | 38% | 8.6 | 2756 |
| 12-17 | 31726 | 23.6 | 37.7 | 60% | 14.1 | 4473 |
| 18-24 | 42162 | 25.4 | 41.8 | 65% | 16.4 | 6915 |
| 25-34 | 73285 | 25.9 | 31.9 | 23% | 6.0 | 4397 |
| 35-49 | 115274 | 23.6 | 32.2 | 36% | 8.6 | 9914 |
| 50-64 | 99841 | 21.2 | 29.8 | 41% | 8.6 | 8586 |
| 65-74 | 40317 | 14.9 | 22.5 | 51% | 7.6 | 3064 |
| ≥75 | 43495 | 9.3 | 16.2 | 74% | 6.9 | 3001 |

[a] Seroprevalence based on results from Roche N test only.

Percent increase calculated as: ((Jun-Jul seroprevalence / Nov-Dec seroprevalence) – 1) x 100.

Absolute increase calculated as: Jun-Jul seroprevalence – Nov-Dec seroprevalence.

Absolute increase N calculated as: absolute increase % x Geneva population

Seroprevalence estimates for November-December 2020 from previous seroprevalence study [2].

Data on Geneva population available from: https://www.ge.ch/statistique/domaines/01/01_01/tableaux.asp#5

**Table S4. Comparison of education level in sample population and Geneva population**

| Education level | Geneva population No. (%) | Study sample No. (%) |
|---|---|---|
| Mandatory | 98246 (26.6) | 203 (8.1) |
| Secondary | 118125 (32.0) | 818 (32.5) |
| Tertiary | 153334 (41.5) | 1499 (59.5) |

Geneva population data available from: https://www.ge.ch/statistique/domaines/15/15_03/tableaux.asp#1

**Table S5. Comparison of proportion vaccinated in sample population and Geneva population**

| | Study sample | | Geneva population | |
|---|---|---|---|---|
| **Age group, years** | **Individuals N** | **Vaccinated N (%)** | **Individuals N** | **Vaccinated N (%)** |
| 0-9 | 328 | 0 (0) | 52912 | 13 (0.02) |
| 10-19 | 444 | 56 (12.6) | 53165 | 7604 (14.3) |
| 20-29 | 306 | 131 (42.8) | 65068 | 27492 (42.3) |
| 30-39 | 423 | 189 (44.7) | 76120 | 37679 (49.5) |
| 40-49 | 558 | 312 (55.9) | 76190 | 47030 (61.7) |
| 50-59 | 505 | 363 (71.9) | 71485 | 49217 (68.9) |
| 60-69 | 304 | 241 (79.3) | 46829 | 35281 (75.3) |
| 70-79 | 236 | 218 (92.4) | 36581 | 30053 (82.2) |
| ≥80 | 81 | 75 (92.6) | 25778 | 21338 (82.7) |

Data on individuals vaccinated with at least 1 dose in study sample up to 4 July, 2021, to match data on individuals vaccinated in the general population of Geneva with at least 1 dose up to July 4, 2021.

Geneva population data available from:
https://www.covid19.admin.ch/en/vaccination/persons/d/demography?geo=GE&demoSum=total&demoAge=minOne

### References


1 Stringhini S, Wisniak A, Piumatti G, *et al.* Seroprevalence of anti-SARS-CoV-2 IgG antibodies in Geneva, Switzerland (SEROCoV-POP): a population-based study. *The Lancet* 2020; **396**: 313–9.

2 Stringhini S, Zaballa M-E, Perez-Saez J, *et al.* Seroprevalence of anti-SARS-CoV-2 antibodies after the second pandemic peak. *The Lancet Infectious Diseases* 2021; **21**: 600–1.

3 Perez-Saez J, Zaballa M-E, Yerly S, *et al.* Persistence of anti-sars-cov-2 antibodies: immunoassay heterogeneity and implications for serosurveillance. *Clinical Microbiology and Infection* DOI:10.1016/j.cmi.2021.06.040.

4 L'Huillier AG, Meyer B, Andrey DO, *et al.* Antibody persistence in the first 6 months following SARS-CoV-2 infection among hospital workers: a prospective longitudinal study. *Clinical Microbiology and Infection* 2021; **27**: 784.e1-784.e8.

5 Baden LR, El Sahly HM, Essink B, *et al.* Efficacy and Safety of the mRNA-1273 SARS-CoV-2 Vaccine. *N Engl J Med* 2020; **384**: 403–16.

6 Polack FP, Thomas SJ, Kitchin N, *et al.* Safety and Efficacy of the BNT162b2 mRNA Covid-19 Vaccine. *N Engl J Med* 2020; **383**: 2603–15.

7 Gelman A, Carpenter B. Bayesian analysis of tests with unknown specificity and sensitivity. *Journal of the Royal Statistical Society: Series C (Applied Statistics)* 2020; **69**: 1269–83.

8 Muench P, Jochum S, Wenderoth V, *et al.* Development and validation of the Elecsys Anti-SARS-CoV-2 immunoassay as a highly specific tool for determining past exposure to SARS-CoV-2. *Journal of Clinical Microbiology* 2020; **58**: e01694-20.

9 Ainsworth M, Andersson M, Auckland K, *et al.* Performance characteristics of five immunoassays for SARS-CoV-2: a head-to-head benchmark comparison. *The Lancet Infectious Diseases* 2020; **20**: 1390–400.

10 Stan Development Team. Stan Modeling Language Users Guide and Reference Manual. Version 2.21.0. 2019. https://mc-stan.org.

11 Stan Development Team. Rstan: the R interface to Stan. R package version 2.21.2. 2020. https://mc-stan.org.

12 R Core Team. R: A Language and Environment for Statistical Computing. Vienna, Austria, 2021 https://www.R-project.org/.

13 Gabry J. shinystan: Interactive Visual and Numerical Diagnostics and Posterior Analysis for Bayesian Models. R package version 2.5.0. 2018. https://CRAN.R- project.org/ package=shinystan.

STROBE Statement—Checklist of items that should be included in reports of ***cross-sectional studies***

| | Item No | Recommendation | Page No |
|---|---|---|---|
| **Title and abstract** | 1 | (*a*) Indicate the study's design with a commonly used term in the title or the abstract | 1 |
| | | (*b*) Provide in the abstract an informative and balanced summary of what was done and what was found | 2 |
| **Introduction** | | | |
| Background/rationale | 2 | Explain the scientific background and rationale for the investigation being reported | 3 |
| Objectives | 3 | State specific objectives, including any prespecified hypotheses | 3 |
| **Methods** | | | |
| Study design | 4 | Present key elements of study design early in the paper | 3-4 |
| Setting | 5 | Describe the setting, locations, and relevant dates, including periods of recruitment, exposure, follow-up, and data collection | 3-4 |
| Participants | 6 | (*a*) Give the eligibility criteria, and the sources and methods of selection of participants | 3, Addendum S2 |
| Variables | 7 | Clearly define all outcomes, exposures, predictors, potential confounders, and effect modifiers. Give diagnostic criteria, if applicable | 4, Addenda S3-4 |
| Data sources/ measurement | 8* | For each variable of interest, give sources of data and details of methods of assessment (measurement). Describe comparability of assessment methods if there is more than one group | 3-4, Addenda S2-4 |
| Bias | 9 | Describe any efforts to address potential sources of bias | Addenda S2-4 |
| Study size | 10 | Explain how the study size was arrived at | Figure S1 |
| Quantitative variables | 11 | Explain how quantitative variables were handled in the analyses. If applicable, describe which groupings were chosen and why | Addendum S4 |
| Statistical methods | 12 | (*a*) Describe all statistical methods, including those used to control for confounding | Addendum S4 |
| | | (*b*) Describe any methods used to examine subgroups and interactions | Addendum S4 |
| | | (*c*) Explain how missing data were addressed | Addendum S4 |
| | | (*d*) If applicable, describe analytical methods taking account of sampling strategy | Addendum S4 |

| | | | |
|---|---|---|---|
| | | (*e*) Describe any sensitivity analyses | Addendum S4 |
| **Results** | | | |
| Participants | 13* | (a) Report numbers of individuals at each stage of study—eg numbers potentially eligible, examined for eligibility, confirmed eligible, included in the study, completing follow-up, and analysed | Figure S1 |
| | | (b) Give reasons for non-participation at each stage | Figure S1 |
| | | (c) Consider use of a flow diagram | Figure S1 |
| Descriptive data | 14* | (a) Give characteristics of study participants (eg demographic, clinical, social) and information on exposures and potential confounders | 4 |
| | | (b) Indicate number of participants with missing data for each variable of interest | Figure S1 |
| Outcome data | 15* | Report numbers of outcome events or summary measures | 4-5 |
| Main results | 16 | (*a*) Give unadjusted estimates and, if applicable, confounder-adjusted estimates and their precision (eg, 95% confidence interval). Make clear which confounders were adjusted for and why they were included | 4-5 |
| | | (*b*) Report category boundaries when continuous variables were categorized | NA |
| | | (*c*) If relevant, consider translating estimates of relative risk into absolute risk for a meaningful time period | NA |
| Other analyses | 17 | Report other analyses done—eg analyses of subgroups and interactions, and sensitivity analyses | NA |
| **Discussion** | | | |
| Key results | 18 | Summarise key results with reference to study objectives | 5-7 |
| Limitations | 19 | Discuss limitations of the study, taking into account sources of potential bias or imprecision. Discuss both direction and magnitude of any potential bias | 7 |
| Interpretation | 20 | Give a cautious overall interpretation of results considering objectives, limitations, multiplicity of analyses, results from similar studies, and other relevant evidence | 7 |
| Generalisability | 21 | Discuss the generalisability (external validity) of the study results | 7 |
| **Other information** | | | |
| Funding | 22 | Give the source of funding and the role of the funders for the present study and, if applicable, for the original study on which the present article is based | 4 |

*Give information separately for exposed and unexposed groups.

**Note:** An Explanation and Elaboration article discusses each checklist item and gives methodological background and published examples of transparent reporting. The STROBE checklist is best used in conjunction with this article (freely available on the Web sites of PLoS Medicine at http://www.plosmedicine.org/, Annals of Internal Medicine at http://www.annals.org/, and Epidemiology at http://www.epidem.com/). Information on the STROBE Initiative is available at www.strobe-statement.org.
